# Variability in US COVID Mortality, Viral Evolution, and the Emergence of Acquired Social Immune Dysfunction

**DOI:** 10.64898/2026.06.29.26356883

**Authors:** Robert D. Morris

## Abstract

**Background:** The course of a pandemic depends on viral traits, available public health interventions, implementation, and individual behavior, all of which interact and vary over time and space. Understanding COVID-19 therefore requires considering these factors together.

**Methods:** National mortality, state-level mortality per 100,000 population for four representative states, policy stringency, and vaccination coverage were assembled from publicly available sources for February 2020 through September 2022. Viral traits, mutation rates, and vaccine effectiveness were drawn from published systematic reviews and standardized relative to wild-type values.

**Results:** Each state experienced a different worst wave, separated by as much as 18 months, with up to a 24-fold difference in state mortality during major waves. NPI stringency was initially high and responsive to COVID waves, but declined during 2021 in both magnitude and responsiveness as vaccination levels rose, then flattened. Viral evolution first increased transmissibility; as population immunity rose, immune escape increased sharply, particularly with Omicron, which produced similar mortality peaks in all four states and the second largest national peak.

**Conclusions:** COVID-19 did not unfold as a single national pandemic but as regionally divergent epidemics that fragmented public perception and weakened cohesion. At the same time, SARS-CoV-2 evolved traits that reduced the apparent and actual effectiveness of interventions. Omicron brought these processes together: despite producing one of the largest national mortality waves, it elicited little renewed policy activation or booster uptake. I describe this progressive uncoupling of epidemic threat, intervention effectiveness, policy activation, and public compliance as Acquired Social Immune Dysfunction.

## Introduction

The effectiveness of a society’s pandemic response depends on its capacity to perceive a biological threat and mount a coordinated response. That capacity, referred to here as the social immune system, involves three interacting components: available interventions (conditioned on their effectiveness), policies to implement those interventions and public compliance. When these components function together and remain coupled to the threat signal, they can substantially limit epidemic spread and mortality. When they decouple from the threat, mortality accumulates despite the availability of effective tools.

Over two and a half years of the COVID-19 pandemic, the United States experienced a series of major mortality waves. The size and diversity of the country meant the local and regional experience differed dramatically as a function of geography, individual behavior, and local public health response.^1,2^ Simultaneously, the virus was evolving in response to selection pressure exerted by the collective effect of individual immunity and behavior.

The heterogeneity of the public health response to the pandemic varied substantially among states and the overall decline in compliance over time.^3–5^ At the same time, SARS-CoV-2 evolved in ways that had a major impact on the effectiveness of the public health response.^6–8^ This study describes the geographic variability in the impact of COVID, shifts in the genotype and phenotype of the virus, and trends in the public health response over the course of the pandemic. The resulting time series create a framework for evaluating the relative health of our social immune system over time and to examine possible causes for any declines in that response. This analysis is descriptive and interpretive. It uses trends in mortality, policy, vaccination, and viral trait to identify temporal patterns to develop a conceptual model of pandemic response failure, rather than to estimate causal effects. It introduces Acquired Social Immune Dysfunction to describe a progressive loss of coupling between epidemic threat, available interventions, policy activation, and public compliance.

## Methods

This study characterizes the interrelationship among epidemiological impact, public health response, and viral evolution over the course of the COVID-19 pandemic in the United States from February 2020 through September 2022. Data were assembled from publicly available sources across four domains: national mortality, state-level mortality, public health response, and viral characteristics.

### Mortality

National COVID-19 mortality was obtained from Our World in Data,^9^ which compiled data from the World Health Organization and the U.S. Centers for Disease Control and Prevention. Daily death counts were smoothed with a seven-day rolling average.

State-level COVID-19 mortality data were obtained from the New York Times COVID-19 Data Repository (github.com/nytimes/covid-19-data)^10^ which compiled daily cumulative case and death counts from state and local health agencies through March 23, 2023. Daily death counts were derived from cumulative totals, smoothed with a seven-day rolling average, and expressed as rates per 100,000 population using denominators from the 2020 U.S. Census.^11^ Four states were selected to illustrate divergent pandemic experiences: New York, Florida, South Dakota, and California. These states were selected as archetypes of distinct epidemic phenotypes reflecting differences in population density, climate, inbound visitor volume, and public health policy, not as a representative sample.

### Public health response

Vaccination coverage and the Oxford COVID-19 Government Response Stringency Index were obtained from Our World in Data,^9^ drawing on data from the U.S. Centers for Disease Control and Prevention and the Oxford COVID-19 Government Response Tracker maintained by the Blavatnik School of Government, University of Oxford.^12^ The Oxford Stringency Index is interpreted as a partial indicator of formal policy activation rather than a complete measure of public health response. In the United States, where policy and adherence varied substantially across states, institutions, communities, and individuals,^3^ stringency is treated as one visible institutional component of a broader response that also includes vaccination, booster uptake, testing, isolation, masking, risk communication, and public acceptance.

### Viral traits

Three viral traits were used to characterize the prevalent strain of SARS-CoV-2: transmissibility (estimated R0), intrinsic severity (infection fatality rate, IFR), and vaccine effectiveness against infection (VE-infection), used as an imperfect proxy for effectiveness against transmission. Estimates were obtained from published systematic reviews, meta-analyses, and large observational studies for each major variant period.^13–18^ All three traits were standardized relative to wild-type values to facilitate comparison across variant periods. Where the literature provides heterogeneous estimates, values representative of direction and relative magnitude were used.

### Viral genomics

The number of nucleotide mutations relative to the wild-type reference sequence (GenBank NC_045512.2) and the proportion of mutations located within the spike protein coding sequence were obtained from the Nextstrain SARS-CoV-2 genomic database.^19^ Values were converted to median monthly estimates. The spike protein coding sequence constitutes approximately 13% of the 29,903-nucleotide SARS-CoV-2 genome.

### Analytic approach

Viral trait, genomic, and public health response data were organized by variant epoch. Variant eras are used as anchor points summarizing a continuous evolutionary process rather than as discrete biological transitions. State-level mortality data were examined for local maxima within and across variant periods to characterize geographic dispersion. The ratio of the highest to lowest state mortality rate at each local maximum was used as an index of geographic dispersion.

## Results

The national morbidity and mortality time series are shown in Figure 1a. Figure 1b shows COVID-19 mortality per 100,000 population for New York, Florida, California and South Dakota. Each state experienced a different wave as its worst, separated by as much as 18 months. New York’s worst wave was the spring 2020 wild-type surge, reaching nearly 4.8 deaths per 100,000 in April 2020. South Dakota’s worst wave was the Alpha period in December 2020, reaching nearly 3.0 per 100,000. California did not experience its mortality peak until the winter of 2021 and Florida’s worst wave was the Delta period in September 2021, reaching approximately 2.1 per 100,000. Omicron, by contrast, produced moderate and nearly identical mortality across all four states simultaneously — the only wave to do so. To distinguish regionally distinct local maxima in the summer of 2020 and the fall of 2021, they are labeled as Wild Type 2 and Alpha/Sturgis. Table 1 summarizes the state-specific magnitude of these local maxima and their relative magnitudes.

**Figure 1a.**
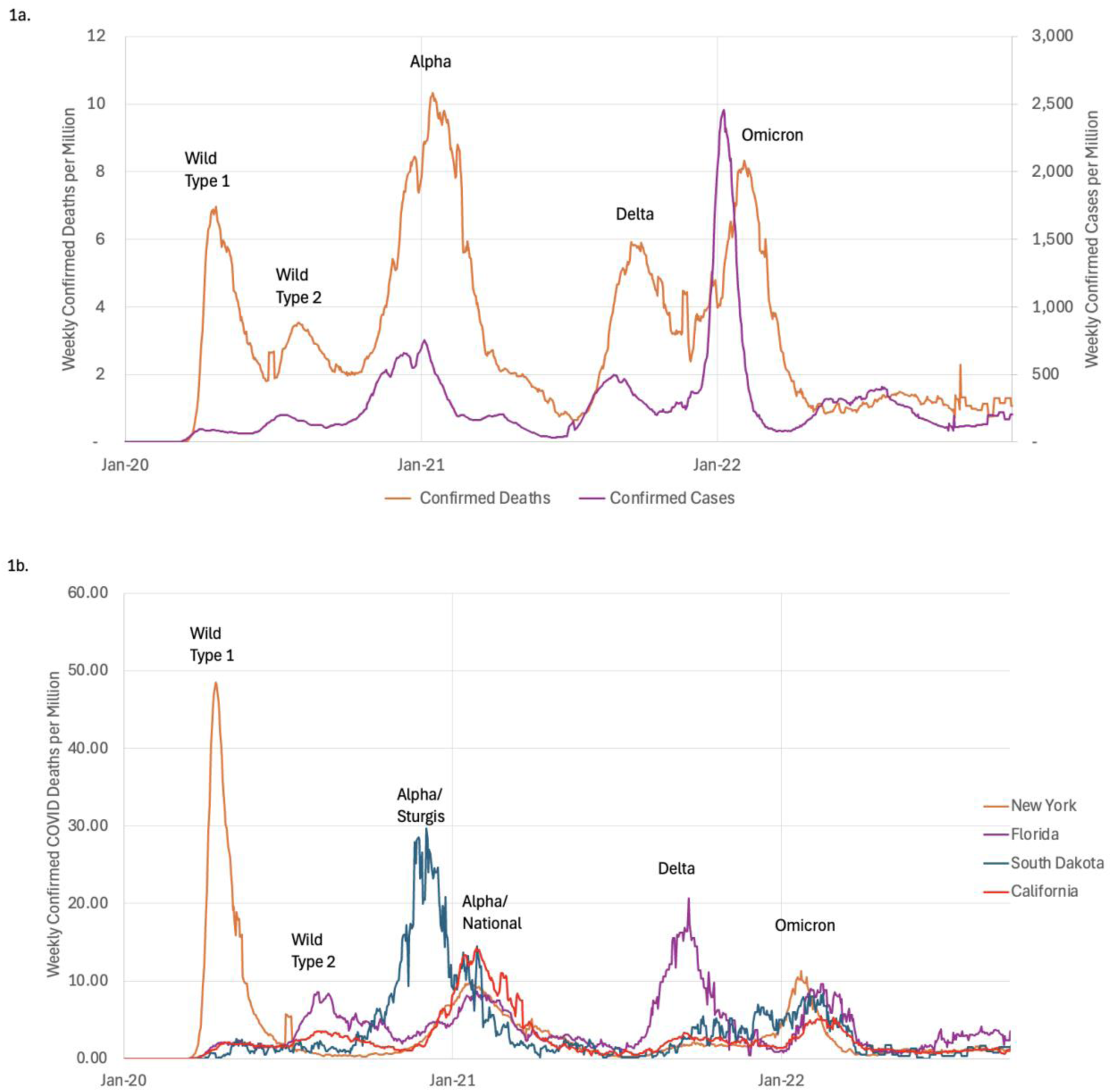
National COVID-19 mortality and confirmed cases, United States, January 2020 – December 2022. Daily confirmed deaths per million (left axis, 7-day rolling average) and weekly confirmed cases per million (right axis) are shown across five variant wave epochs: Wild-type 1, Wild-type 2, Alpha, Delta, and Omicron. **Figure 1b**. COVID-19 mortality by state, February 2020 – September 2022. Weekly COVID-19 mortality rate per 100,000 population for New York, Florida, South Dakota, and California. Vertical lines demarcate the six local mortality maxima identified in the state-level data.

**Table 1.** Peak weekly COVID-19 mortality rate (deaths per 100,000) by state and pandemic surge. The bolded value in each row indicates the state most affected by that surge.

| Surge | New York | Florida | South Dakota | California | Max/Min |
| --- | --- | --- | --- | --- | --- |
| Wild 1 | **4.84** | 0.24 | 0.26 | 0.21 | 23.0× |
| Wild 2 | 0.12 | **0.86** | 0.23 | 0.36 | 7.2× |
| Sturgis | 0.90 | 0.72 | **2.96** | 1.31 | 4.1× |
| Alpha (minus Sturgis) | 0.98 | 0.86 | **1.45** | 1.42 | 1.7× |
| Delta | 0.21 | **2.06** | 0.55 | 0.33 | 9.8× |
| Omicron | **1.13** | 0.96 | 0.84 | 0.54 | 2.1× |

*Until Omicron, every surge had a single dramatic outlier. The Sturgis Motorcycle Rally surge (fall 2020) is shown separately from the Alpha wave because it was a discrete superspreader event preceding the variant's national spread; its removal accounts for the otherwise anomalous uniformity of the Alpha wave. The Max/Min column gives the ratio of the highest to lowest state mortality rate in each surge. Data source: New York Times COVID-19 Data Repository; 2020 U.S. Census denominators.*

Figures 2a and 2b show viral mutation counts and viral traits over the period of interest. There is a steady rise in R_0_ over the course of the pandemic with the sharpest rise between the Alpha and Delta variants. At the same time, vaccine effectiveness dropped steadily with particularly sharp drops for Delta and Omicron. Disease severity rose for both Alpha and Delta variants before dropping more than 50% between Delta and Omicron.

**Figure 2a.**
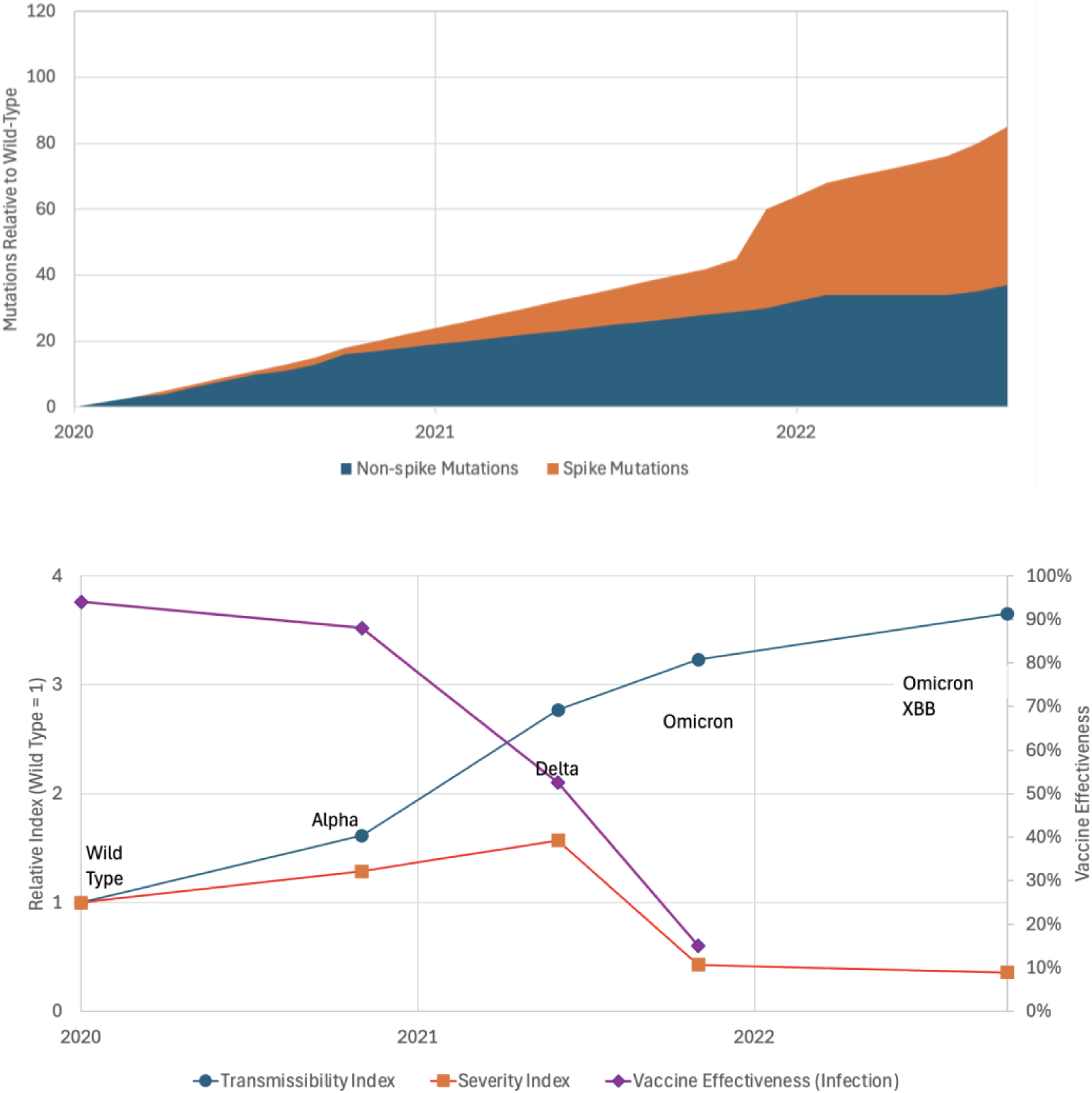
Directional changes in key SARS-CoV-2 viral traits across major variant eras. Relative transmissibility (R0), intrinsic severity (IFR), and vaccine effectiveness against infection are shown for the wild-type, Alpha, Delta, and Omicron eras, normalized to wild-type values (wild-type = 1.0 for R0 and IFR; wild-type vaccine effectiveness shown on right axis). Values represent central estimates from the published literature and are intended to convey relative magnitude and direction rather than precise parameters. Intrinsic severity reflects risk in immunologically naïve or minimally immune populations. **Figure 2b**. Trends in SARS-CoV-2 genomic mutations relative to the wild-type reference sequence (GenBank NC_045512.2), January 2020 – August 2022. Monthly median total mutations are shown as stacked areas representing non-spike (blue) and spike (orange) mutations. Data source: Nextstrain SARS-CoV-2 genomic database.

These changes are mirrored in the nature and number of mutations in the viral genome. The total number of mutations rose at a relatively constant rate from the wild-type through to the Delta variant. During the first 10 months, those mutations were spread relatively evenly across the genome with the gene for the spike protein, which constitutes 13% of the viral genome,^20^ accounting for 11% of the mutations in the Alpha variant. That proportion rose from Alpha to 30% of the Delta mutations and 50% of the Omicron mutations. The leap from Delta to Omicron also demonstrated a sharp increase in the total number of mutations.

Figure 3 depicts the public health response against the national COVID mortality time series. The stringency index for NPIs is shown in the green curve, showing relatively strict interventions until the Alpha wave began to decline in the winter of 2021. It then dropped steadily over the next two years other than a slight uptick in response to the Delta wave. Notably, there was no rise in stringency in response to the Omicron wave.

**Figure 3.**
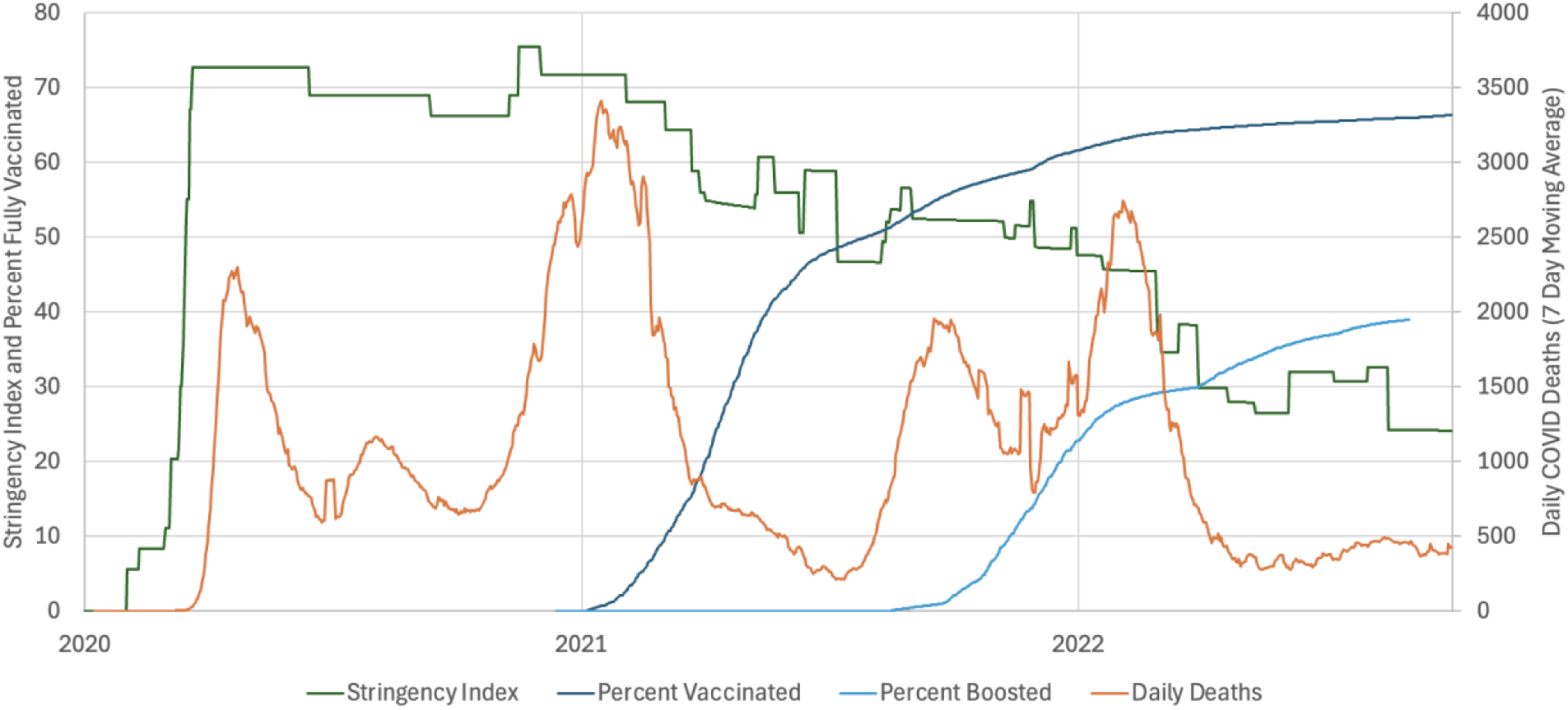
National public health response and COVID-19 mortality, United States, January 2020 – December 2022. The Oxford COVID-19 Government Response Stringency Index (green, left axis), percentage of the population fully vaccinated (dark blue, left axis), and percentage boosted (light blue, left axis) are shown alongside daily COVID-19 deaths (orange, right axis, 7-day rolling average). Data sources: Our World in Data; Oxford COVID-19 Government Response Tracker.

Vaccination rates, shown as the percentage of the population that had received the full vaccination course of two vaccines, rose sharply in the first half of 2021 as the Alpha wave was declining, hitting 50% as the Delta wave arrived and rising to 65% during the Omicron wave. The first booster was released during the Alpha wave and usage quickly rose to 20% and had passed 25% before the Delta wave began its sharp rise. By the time it reached 30%, the Delta wave was already fading. The final rise in booster uptake reached only 35% and occurred after the Delta wave had faded.

Table 2 summarizes the viral traits and response measures associated with each major wave.

**Table 2.** Viral traits and public health response across the major SARS-CoV-2 variant waves in the United States. Deaths are approximate totals attributed to each wave. Relative R0 and relative IFR are expressed relative to the wild-type strain (wild type = 1). The spike mutation ratio is the number of spike-region mutations as a proportion of total mutations, with all counts taken relative to the wild-type genome. Stringency values are the maximum and minimum of the Oxford Government Response Stringency Index during each wave (max/min). Vaccine and booster values are coverage before and at the end of each wave (pre/post).

|  | Wild Type 1 | Wild Type 2 | Alpha | Delta | Omicron |
| --- | --- | --- | --- | --- | --- |
| **Start** | 2/1/20 | 6/25/20 | 10/15/20 | 7/11/21 | 11/29/21 |
| **End** | 6/25/20 | 10/15/20 | 7/11/21 | 11/20/21 | 6/22/22 |
| **Deaths** | 123,000 | 97,000 | 383,000 | 165,000 | 234,000 |
| ***Viral traits*** |  |  |  |  |  |
| **Relative $R_0$** | 1 | 1? | 1.6 | 2.7 | 3.2 |
| **Relative IFR** | 1 | 1? | 1.2 | 1.5 | 0.5 |
| **Total Mutations** | - | ? | 16 | 25 | 60 |
| **% Spike Mutations** | - | ? | 11% | 31% | 50% |
| ***Public health response*** |  |  |  |  |  |
| **Stringency (max/min)** | 73/69 | 69/66 | 75/50 | 57/48 | 51/26 |
| **Vaccine (pre/post)** |  |  |  | 50/60 | 60/65 |
| **Booster (pre/post)** |  |  |  | 0/20 | 20/30 |

## Discussion

Overall, the local severity of the pandemic in a particular area at a given time is a consequence of the interplay the traits of the virus and three characteristics of that location: geographic factors, individual factors, and public health interventions. Geographic factors include population density, demographics, visitor influx and weather, which are highly time variant. Individual factors include immunity and behavior. Public health interventions seek to limit the impact of all these factors on the spread of disease. We can see each of these factors at work in these data.

Nationally we see four clear mortality peaks with a smaller fifth peak in the summer of 2020. Morbidity data demonstrate first the variable reliability of these data, particularly during the initial wave but also indicate a dramatic reduction if case fatality rates during Omicron. Figure 1b demonstrates the dramatic effect of geographic factors. The individual state time series tell a story that is invisible in the national aggregate data. These four states each experienced their worst episode of the pandemic at a different time.

New York’s catastrophic spring 2020 surge reflected the intersection of high population density, extensive inbound international travel, cold-weather indoor crowding, and a complete absence of prior population immunity or pharmaceutical countermeasures. The New York peak weekly mortality during that initial surge was 21, 23, and 19 times that of California, Florida, and South Dakota respectively. The New York experience set the national template for rapid, universal, and stringent NPI response at a moment when much of the rest of the country had not yet experienced significant mortality.

Due at least in part to those strict NPIs, the pandemic experience differed radically among states, with the other states experience minor mortality peaks compared to New York. To some extent, those public health measures may have been a victim of their own success. Effective prevention measures create the unfortunate illusion that they were unnecessary.

California, with a mild winter climate and implementation of NPIs before the first surge in COVID cases had a peak mortality rate during the first wave that was one twentieth of New York’s. This divergence in local experience may help explain why arguments minimizing COVID-19 severity gained early traction in California. Several California-based researchers published prominent opinion pieces^21–23^ suggesting that infection severity was being overestimated, and later conducted a seroprevalence study^24,25^ whose non-standard recruitment and analytic methods yielded an unusually low IFR estimate. That estimate became an important source of support for minimalist public health strategies.

Florida experienced the brunt of the smaller secondary peak, which followed a fundamentally different seasonal pattern, with major peaks in the summers of 2020 and 2021 as heat drove people indoors to create the Sun Belt analog of the Northeast’s winter transmission dynamics. This phase shift between COVID peaks in southern and northern states helps to explain the resistance to policies heavily influenced by the northern experience.

Despite having far more severe winter weather than New York, South Dakota, which ranks 47^th^ for international visitors^26^ and 46^th^ for population density^11^ among US states, experienced a dramatically different pandemic from the other four states. Its massive surge in mortality occurred in the fall of 2020, prior to the emergence of the Alpha variant, as the other three states were experiencing a minimum. This peak represents the first dramatic local failure of the social immune system related to the Sturgis Motorcycle Rally, which created a massive visitor influx, a surge in population density, and a lack of compliance with NPIs.

The second dramatic failure of the local social immune system occurred during the Delta wave in the fall of 2021, which was only prominent among these states in Florida. In July of 2021, Ron DeSantis issued an executive order banning school mask mandates.^27^ That September, the Delta wave peaked in Florida earlier and higher than the other states at a mortality rate almost ten times New York. Note that the ban on school masking reflects a broader push against public health interventions in Florida and is unlikely to be exclusively responsible for that surge.

### The national public health response

The national public health response, as shown in Figure 3, exhibits three distinct phases: an NPI-dominant phase in 2020, a transitional phase in 2021 marked by the introduction of vaccination, and an attenuated response phase in late 2021 and into 2022.

During the NPI phase, stringency rose rapidly to a peak of 73 in spring 2020 and remained elevated through the year, declining only modestly to 66 before the Alpha wave arrived. This sustained response coincided with the smallest mortality peak of the pandemic, though the causal inference is complicated by the geographic concentration of that wave. By the time Alpha emerged, the social context in which NPIs operated had changed. The severity of COVID-19 had been called into question, the value of NPIs had become politically contentious, and pandemic fatigue had begun to erode voluntary adherence. Stringency peaked again at 75 during Alpha but the same formal index likely translated into less effective behavioral change than it had in 2020.

The transitional phase began with vaccine authorization in late 2020 and accelerated through 2021. As vaccination coverage rose from near zero to roughly 50% by the Delta wave, formal NPI stringency declined from its Alpha peak to 57, then 48. This was not straightforward disengagement. Delta’s severity sustained a sense of threat, and the social immune response had shifted instruments: vaccination and boosting replaced NPIs as the primary expression of collective response. Booster uptake rose from zero to roughly 20% during and after the Delta wave. The system was still responding, but through a different channel.

The attenuated phase arrived with Omicron. Stringency did not rise to meet the wave; it fell throughout, from 51 to 26, inverting the pattern seen in every prior wave. Vaccination coverage rose only modestly to 65%, and booster uptake, despite ample supply and months of availability, plateaued below 40%. Two thirds of the US population remained unboosted as the second-deadliest wave of the pandemic peaked. The smaller Delta threat had drawn a larger response than the larger Omicron one. This inversion is the empirical signature of ASID.

### Viral evolution and the degradation of the response signal

These shifts in the public health response did not occur in isolation. The virus was evolving in ways that made the response progressively harder to sustain.

During the NPI phase, selective pressure favored transmissibility. Alpha and Delta emerged with relative R0 values of 1.6 and 2.7 respectively, each wave more transmissible than the last. More transmissible variants are inherently harder to suppress through behavioral means, meaning that each successive NPI response was working against a more demanding biological target even as the behavioral response was weakening. At the same time, mutations during this phase were distributed relatively evenly across the genome.

As the transitional phase brought widespread vaccination, the selective landscape shifted. Spike-directed immunity became the dominant constraint on viral replication, and the genomic record reflects this. The proportion of mutations in the spike protein rose from 11% at the start of Alpha to 30% at the start of Delta, three times the gene’s proportional share. This is the molecular signature of rising selection pressure for immune escape, driven by the same vaccination campaign that was simultaneously serving as the social immune system’s primary response instrument.

Omicron represented a discontinuity rather than an extrapolation. Spike mutations rose to 50% of all mutations, accompanied by a sharp increase in total mutation burden. Transmissibility reached a relative R_0_ of approximately 3.2 while intrinsic severity fell to roughly half the wild-type IFR. The resulting phenotype was precisely suited to exploit the attenuated response phase: highly transmissible through immune populations, producing infections visible enough to undermine vaccine confidence, but with per-infection lethality low enough to weaken the threat signal the response had been calibrated to detect. Vaccine effectiveness against infection collapsed while protection against severe disease was substantially preserved, a distinction the response signal could not easily communicate and the public could not easily perceive.

### Omicron, ASID, and converging causes

The Omicron wave is where the geographic, epistemic, and biological threads converge.

Geographically, Omicron was the first wave to strike all four states simultaneously and at comparable magnitude. Every prior surge had a major outlier — a state experiencing catastrophe while the others were near baseline (Table 1). Omicron’s Max/Min ratio of 2.1 was the lowest of any wave, approaching uniformity. The synchronicity of these relatively small waves, made Omicron the second largest wave of the entire pandemic. This convergence was not the result of reduced viral danger. It was the result of immune escape overriding the geographic, seasonal, and behavioral factors that had previously stratified epidemic experience. For the first time, the national mortality curve reflected something genuinely shared. But the social immune system that might have responded to a shared threat had been systematically eroded by 18 months of surges that were never shared, each one reinforcing locally valid but nationally incompatible beliefs about the pandemic’s nature and severity.

The epistemic erosion had begun early. The California-based seroprevalence study, conducted at a moment when that state’s mortality was one-twentieth of New York’s, provided early scientific cover for minimizing the pandemic’s severity. Published during the NPI phase, it made the argument for a far lower IFR credible to populations whose own experience seemed to support it but incredible to populations who had watched thousands die in a matter of weeks. The asymmetry was not one of rationality but of geography. Two years later, the institutional prohibition on school masking in Florida during the Delta peak represented a different mode of epistemic failure: not a researcher misreading local data but a government actively dismantling an available intervention during an active surge, accelerating the divergence between formal policy and effective response that the Oxford Stringency Index could measure but not capture. The fact that Florida, with summer surges, was out of phase with the rest of the country helps explain its resistance to NPIs.

The viral evolution documented in the traits and genomic data contributed to the confusion. Evolution did not select for confusion directly. It selected for traits that favored replication in an environment where immunity constrained transmission, and those traits incidentally generated the observations on which denialism drew. By degrading vaccine effectiveness and causing breakthrough infections, Omicron produced a true signal that supported a false inference that vaccines had failed. By reducing per-infection severity while maintaining lethal aggregate burden, it weakened the threat signal the response had been calibrated to detect. The same evolutionary move that escaped the biological immune system’s recognition at the spike partially escaped the social immune system’s threat detection.

Acquired Social Immune Dysfunction is the name for what these three forces produced in combination. The social immune system comprises policy activation, intervention effectiveness, and public compliance. Across the pandemic arc these three components decoupled from the threat signal in sequence: compliance first, through geographic fragmentation and the perception that the national response was calibrated to someone else’s emergency; effectiveness second, as viral evolution made NPIs and then vaccines progressively less effective against infection; and policy last, as institutional will collapsed under the accumulated weight of contested severity, visible breakthrough infection, and two years of incommensurable regional experience. By the time Omicron arrived as the first genuinely universal epidemic, there was no coordinated response left to activate. The dysfunction was not a failure of any single institution or individual. It was the convergent product of a geographically fragmented pandemic, a virus that evolved to degrade the signals sustaining collective response, and an epistemic environment in which that degradation was amplified into national narratives of institutional failure and overreach. Understanding ASID as the interaction of these three forces, rather than as the consequence of any one of them, is a prerequisite for designing social immune systems capable of functioning under the conditions the next pandemic will impose.

## Limitations

This analysis is descriptive and interpretive. It uses quantitative epidemiologic and virologic measures to characterize a qualitative relationship between viral evolution, epidemiologic impact, and public health response, and does not estimate causal effects or treat successive variant waves as exchangeable observations. Each wave occurred in a changing context of population immunity, treatment capacity, viral adaptation, public trust, institutional fatigue, and political conflict that cannot be fully disentangled.

Estimates of transmissibility, intrinsic severity, and vaccine effectiveness vary by study design, population, and time period. The values in Table 2 represent direction and relative magnitude rather than definitive parameters. Variant eras are simplified anchors for a continuous evolutionary process. The four states examined in Figure 1b were selected to illustrate distinct epidemic phenotypes and do not constitute a representative sample; the broader geographic pattern they exemplify is consistent with regional aggregate data but is examined in full elsewhere.

Policy stringency is an imperfect proxy for social immune activation, particularly in the United States, where policy and adherence varied substantially across states, institutions, communities, and individuals. It is interpreted here as one indicator of formal policy activation, alongside vaccination and booster uptake. The booster plateau cannot be attributed to any single cause. The U.S. focus limits generalizability to other national contexts.

These limitations do not eliminate the central pattern motivating this analysis. The divergence between epidemiologic burden and social immune activation across five waves, and its resolution in the absence of any measurable response during the second-deadliest wave of the pandemic, is visible in the data regardless of the precise values assigned to any individual parameter.

## Conclusions

From the moment it began, the COVID-19 pandemic sent confusing signals about its severity and the need for public health interventions. Strict intervention policies driven by early worst case signals seemed like overkill for regions not yet affected and with different risk profiles. The result was not a single, coherent pandemic experience, but rather multiple pandemics depending on time and location.

At the same time, SARS-CoV-2 was evolving within a changing landscape of public health interventions and population immunity. Early increases in transmissibility were met with aggressive, though increasingly unstable, social immune activation. Subsequent evolution favored spike-directed immune escape under vaccination pressure, producing a variant with reduced intrinsic severity but extreme transmissibility and evasion of vaccine-induced protection against infection.

The public health response remained disproportionately coupled to visible per-infection severity rather than to transmission dynamics and absolute mortality burden. As Omicron reduced the former while sustaining the latter, the response signal weakened and then disappeared. The Omicron wave illustrates the terminal expression of ASID: a failure of social immune activation despite ongoing population-level threat, in a population that had lost the shared epidemic experience on which coordinated response depends.

Three forces drove this outcome and reinforced one another. Geographic heterogeneity ensured that no single national narrative of the pandemic was locally accurate everywhere, seeding the incommensurable regional beliefs that made national coordination structurally difficult from the outset. Viral evolution progressively degraded the signals sustaining collective response, first by increasing transmissibility beyond what politically and socially sustainable behavioral interventions could reliably suppress, then by generating immune escape that made both NPIs and vaccines appear to fail for reasons indistinguishable from the claims of those arguing they had never worked. And the amplification of scientific misinformation through conventional and social media converted locally valid skepticism into national narratives of institutional failure, foreclosing the possibility of renewed consensus when the evidence might otherwise have supported it.^28^

Recognizing pandemic response as a co-evolutionary process in which the pathogen can degrade the host population’s capacity to perceive and respond to it reframes preparedness as a problem not only of intervention capacity but of signal integrity. A social immune system that cannot accurately read the threat it faces cannot respond proportionately to it, regardless of the tools available. Furthermore, if the effector component of the feedback loop is dysfunctional, because interventions are ineffective, public health authorities do not implement them, or the population is not compliant, the entire system breaks down. Designing systems capable of maintaining that signal integrity and appropriate response under conditions of geographic heterogeneity, viral evolution, and epistemic disruption is the preparedness challenge the next pandemic will impose.

## Data Availability

All data used in this analysis are publicly available. National COVID-19 mortality, vaccination coverage, and policy stringency data are available from Our World in Data (ourworldindata.org/coronavirus). State-level mortality data are available from the New York Times COVID-19 Data Repository (github.com/nytimes/covid-19-data). Genomic mutation data are available from the Nextstrain SARS-CoV-2 database (nextstrain.org/ncov). The SARS-CoV-2 reference genome is available from NCBI GenBank (accession NC_045512.2). Viral trait estimates were derived from published literature cited in the references. No new data were generated for this study.

https://www.bsg.ox.ac.uk/research/covid-19-government-response-tracker

https://github.com/nytimes/covid-19-data

https://www.census.gov/programs-surveys/decennial-census/decade/2020/2020-census-main.html

https://nextstrain.org/ncov

https://www.ncbi.nlm.nih.gov/nuccore/NC_045512.2

## References

1. Jackson, S. L. et al. Spatial Disparities of COVID-19 Cases and Fatalities in United States Counties. International Journal of Environmental Research and Public Health 18, 8259 (2021).

2. Rodriguez, C. G., Gadarian, S. K., Goodman, S. W. & Pepinsky, T. B. Morbid Polarization: Exposure to COVID-19 and Partisan Disagreement about Pandemic Response. Political Psychology 43, 1169–1189 (2022).

3. Koziol, J. A. & Schnitzer, J. E. State Government Policy Responses to the COVID-19 Pandemic in the United States 2020-2022: Concordant Heterogeneity? Medical Research Archives 11, (2023).

4. Agyapon-Ntra, K. & McSharry, P. E. A global analysis of the effectiveness of policy responses to COVID-19. Sci Rep 13, 5629 (2023).

5. Jørgensen, F., Bor, A., Rasmussen, M. S., Lindholt, M. F. & Petersen, M. B. Pandemic fatigue fueled political discontent during the COVID-19 pandemic. Proceedings of the National Academy of Sciences 119, e2201266119 (2022).

6. van Oosterhout, C., Hall, N., Ly, H. & Tyler, K. M. COVID-19 evolution during the pandemic – Implications of new SARS-CoV-2 variants on disease control and public health policies. Virulence 12, 507–508 (2021).

7. van Oosterhout, C. et al. COVID-19 adaptive evolution during the pandemic – Implications of new SARS-CoV-2 variants on public health policies. Virulence 12, 2013–2016 (2021).

8. Angius, F. et al. SARS-CoV-2 Evolution: Implications for Diagnosis, Treatment, Vaccine Effectiveness and Development. Vaccines 13, 17 (2025).

9. Mathieu, E., et al. COVID-19 Pandemic. Our World in Data https://ourworldindata.org/coronavirus (2024).

10. nytimes/covid-19-data. The New York Times (2026).

11. US Census Bureau. Population Density of the 50 States, the District of Columbia, and Puerto Rico: 1910 to 2020. New York https://www2.census.gov/programs-surveys/decennial/2020/data/apportionment/population-density-data-table.pdf (1910).

12. Hale, T. et al. A global panel database of pandemic policies (Oxford COVID-19 Government Response Tracker). Nat Hum Behav 5, 529–538 (2021).

13. Liu, Y. & Rocklöv, J. The reproductive number of the Delta variant of SARS-CoV-2 is far higher compared to the ancestral SARS-CoV-2 virus. Journal of Travel Medicine 10.1093/jtm/taab124 (2021) doi:10.1093/jtm/taab124.

14. Liu, Y. & Rocklöv, J. The effective reproductive number of the Omicron variant of SARS-CoV-2 is several times relative to Delta. J Travel Med 29, taac037 (2022).

15. Meyerowitz-Katz, G. & Merone, L. A systematic review and meta-analysis of published research data on COVID-19 infection fatality rates. International Journal of Infectious Diseases 101, 138–148 (2020).

16. Xia, Q. et al. Case fatality rates of COVID-19 during epidemic periods of variants of concern: A meta-analysis by continents. International Journal of Infectious Diseases 141, 106950 (2024).

17. Mohammed, H. et al. A Systematic Review and Meta-Analysis on the Real-World Effectiveness of COVID-19 Vaccines against Infection, Symptomatic and Severe COVID-19 Disease Caused by the Omicron Variant (B.1.1.529). Vaccines 11, 224 (2023).

18. Song, S., Madewell, Z. J., Liu, M., Longini, I. M. & Yang, Y. Effectiveness of SARS-CoV-2 vaccines against Omicron infection and severe events: a systematic review and meta-analysis of test-negative design studies. Front. Public Health 11, (2023).

19. Nextstrain Team. Genomic epidemiology of SARS-CoV-2. https://nextstrain.org/ncov (2025).

20. Wu, F. et al. A new coronavirus associated with human respiratory disease in China. Nature 579, 265–269 (2020).

21. Sood, N. Opinion | It’s Dangerous to Test Only the Sick. Wall Street Journal (2020).

22. Bendavid, E. & Bhattacharya, J. Opinion | Is the Coronavirus as Deadly as They Say? Wall Street Journal (2020).

23. Ioannidis, J. P. A. A fiasco in the making? As the coronavirus pandemic takes hold, we are making decisions without reliable data. STAT https://www.statnews.com/2020/03/17/a-fiasco-in-the-making-as-the-coronavirus-pandemic-takes-hold-we-are-making-decisions-without-reliable-data/ (2020).

24. Bendavid, E., et al. COVID-19 Antibody Seroprevalence in Santa Clara County, California. http://medrxiv.org/lookup/doi/10.1101/2020.04.14.20062463(2020) doi:10.1101/2020.04.14.20062463.

25. Bendavid, E. et al. COVID-19 antibody seroprevalence in Santa Clara County, California. International Journal of Epidemiology 50, 410–419 (2021).

26. US NTTO. Overseas Visitor Impact on State Economies | 2024. https://www.trade.gov/travel-and-tourism-reports (2024).

27. Governor DeSantis Issues an Executive Order Ensuring Parents’ Freedom to Choose | Executive Office of the Governor.

28. https://www.flgov.com/eog/news/press/2021/governor-desantis-issues-executive-order-ensuring-parents-freedom-choose.

28. Morris, R. D. How denialist amplification spread COVID misinformation and undermined the credibility of public health science. Journal of Public Health Policy 10.1057/s41271-023-00451-4 (2024) doi:10.1057/s41271-023-00451-4.

